# Investigating the Genetics and Antibiotic Resistance of Methicillin Resistant *Staphylococcus aureus* in Biological Samples from Hospitalized Patients

**DOI:** 10.1101/2025.04.11.25325585

**Authors:** Sita Neupane, Aastha Acharya, Samiran Subedi

## Abstract

**Background:** *Staphylococcus aureus* is a gram-positive bacterium that can cause various diseases and infections. Penicillin and methicillin are examples of β-lactam antibiotics, the first line of defense against *Staphylococcus aureus* infections. Methicillin-Resistant *Staphylococcus aureus* (MRSA) is still one of the leading causes of hospital-acquired infections associated with morbidity, mortality, and cost. MRSA can be hospital-acquired (HA-MRSA) or community-associated (CA-MRSA) infections. The main objective of this study is to screen MRSA among HA-MRSA to determine the prevalence of antibiotic susceptibility patterns of MRSA among patients. Furthermore, we identify the *mecA* gene, which produces a penicillin-binding protein (PBP2a) with a low affinity for β-lactam antibiotics.

**Methods:** This study was done on the patients of Kathmandu Model Hospital, Nepal, and the samples were processed at the Microbiology laboratory of Kathmandu Model Hospital. Data analyses were done from Microsoft Excel and GraphPad Prism. DNA extraction was done from the classical CTAB method with minor modifications, and *mecA* gene-specific primers were used to detect the gene in the samples.

**Results:** Out of 4383 samples, 848 (21.00%) samples have growth, and 190(22.4%) were Staphylococcus aureus. Among *Staphylococcus aureus* 52 (27.36%) were Methicillin resistant *Staphylococcus aures*. Antibiotic susceptibility tests were done to characterize MRSA isolates. Most of the isolates were resistant to Amikacin (69.25%), followed by Ampicillin (53.8%), Chloramphenicol (78.84%), Cotrimoxazole (53.8%), Gentamycin (67.3%), Ofloxacin (15.39%), Erythromycin (71.15%) Vancomycin and Teicoplanin (3.84%). In our study, 50 (96.15%) out of 52 MRSA showed the *mecA* gene, while 3.85% showed the absence of the *mecA* gene.

**Conclusions:** The frequency of MRSA infections in HA-MRSA was comparatively high, with a greater abundance of the *mecA* gene that confers the resistance. Regular surveillance of HA-MRSA and genetic profiling of the mecA gene are essential for reducing MRSA infection.

## 1. Introduction

According to the 2014 WHO Global Health Report on antimicrobial resistance in all WHO regions, Methicillin-Resistant *Staphylococcus aureus* (MRSA) prevalence of infections was above 20 % and increased risk of death and associated health care costs [1] (Organization WHO,2014 and Lee BY et al. 2013). MRSA is a worldwide problem not localized to any geographic area [2]. Hospital-acquired MRSA is the most common cause of hospital-acquired infections [3,4]. The most often reported invasive MRSA-related illnesses are septic shock (56%), pneumonia (32%), endocarditis (19%), bacteremia (10%), and cellulitis (6%) [5]. In general, the frequency of infections caused by MRSA has increased in the last decade [6]. Antimicrobial resistance (AMR), which is currently responsible for roughly 5 million deaths annually, has been made worse by the inability to implement infection prevention practices completely and the sharp rise in antibiotic use [7]. Several genetic factors contribute to the multi-drug resistance in HA-MRSA. HA-MRSA has the *SCCmec* gene cassettes that include a *mecA* gene. The *mecA* gene encodes for an alternative penicillin-binding protein 2a (PBP-2a) with low affinity to β-lactams [8].

When β-lactams block native PBPs, the *mecA* gene is necessary for cell wall production. Even within the same species, isolates carrying the same *mecA* gene frequently displayed varying levels of resistance, suggesting that strain-specific factors may be necessary for manifesting methicillin resistance [9].

The main purpose of this research is to detect the presence of the *mecA* gene among the collected samples. Since the horizontal gene transfer of the *mec* gene family has been found to increase the prevalence of MRSA in the community, this study aims to provide general information on how predominant this gene is. This study also seeks to evaluate the efficacy of the disk diffusion test in detecting methicillin-resistant *Staphylococcus aureus*.

## 2. Methods

### 2.1 Sampling

This study takes clinical samples from different biological specimens such as pus, wound, swab, blood, urine, sputum, tissue, nasal swab, and ET. The samples were collected with a sterile swab in Kathmandu Model Hospital. A total of 4285 samples were collected and cultured on nutrient agar. For molecular analysis, MRSA isolates were transported to the Central Department of Biotechnology by taking all the Biosafety and Biosecurity measures according to the guidelines published by National Public Health Laboratory.

### 2.2. Isolation and Identification of *Staphylococcus aureus*

Clinical specimens were cultured in Nutrient Agar and then sub-cultured in 5% sheep blood agar and mannitol salt agar under sterile conditions. The plates were incubated at 37°C under aerobic conditions. Bacterial isolates with golden yellow color on MSA were further analyzed via Gram’s staining, catalase test, slide, and tube coagulase test. Isolates exhibiting the characteristics and properties of *Staphylococcus aureus* were further sub-cultured in blood agar for confirmation.

### 2.3. Antibiotic susceptibility testing

The modified Kirby-Bauer disk diffusion method was used to perform antibiotic susceptibility testing on Mueller Hinton Agar, following the guidelines set by the Clinical Laboratory and Standard Institute [10]. The commercial antibiotics discs and concentrations used were Amikacin(25µg), Ampicillin(10µg), Penicillin G (30µg), Gentamicin(30µg), Cotrimoxazole (25µg), cefoxitin (30µg), Erythromycin (30µg), Ofloxacin, Chloramphenicol, Teicoplanin, Linezolid and vancomycin (30µg). A bacterial suspension equivalent to 0.5 McFarland turbidity standard was prepared for inoculation. The plates were incubated at 37°C for 24 hours in Mueller-Hinton agar (MHA) supplemented with 2% NaCl. An inhibition zone diameter of each antimicrobial was then measured and interpreted as resistant (R), intermediate (I), and sensitive (S) according to CLSI guidelines. MRSA-positive strains were confirmed by those *S. aureus* strains that were resistant to cefoxitin and had a zone size < 21mm.

### 2.4. Extraction of DNA

DNA was extracted by CTAB-the boiling method, in which the pellet of loopful bacteria was resuspended in a TE buffer, followed by adding lysozyme to the mixture. Then proteinase K and 30ul of SDS were added after brief incubation until the suspension became clear. Next, preheated CTAB/NaCl (65°C) was added to the suspension. Then an equal volume of phenol/chloroform/isoamyl alcohol (25:24:1) was added to collect the upper aqueous phase of DNA. To the collected supernatant, isopropanol was added, and DNA was precipitated by adding ethanol to the mixture.

### 2.5. Amplification of *mecA* gene from *Staphylococcus aureus*

For the amplification of the *mecA* gene from *Staphylococcus aureus,* crude lysates were utilized as a DNA template. The primers for this study were used by Vatansever et al., 2016 [14]: Forward Primer (*mecA*PF1) 5’-ACTGCTATCCACCCTCAAAC-3’ and reverse primer (*mecA* PR1) 5’-CTGGTGAAGTTGTAATCTGG-3’. DNA amplification was done in a 10µl of the reaction mixture with 5 μl of master mix, 1 μl of each forward and reverse primer, 1 μl of DNA, and 2 μl of nuclease-free water. The PCR conditions are initial denaturation at 95°C for 120 seconds, denaturation at 95°C for 30 seconds, annealing at 56.2°C for 30 seconds, extension at 72°C for 20 seconds, and 29 amplification cycles at 72°C for 5 minutes. After PCR, the amplicon was analyzed by running the samples in 1.5% agarose gel stained with ethidium bromide. The resulting amplicon length 163-bp was confirmed as a positive sample for the *mecA* gene.

### 2.6. Quality control

The inoculation and culture were carried out using aseptic techniques to ensure contamination-free conditions. The media were checked for the growth of pure cultures of microorganisms. The *S. aureus* ATCC 25923 species was used as a control organism for identification (using gram staining and culture) and antibiotic susceptibility testing. The Mueller-Hinton agar (MHA) thickness was kept at 4 mm, and the pH was maintained between 7.2-7.4. A control smear was stained whenever a new batch of stains was prepared to ensure proper staining reactions. All procedures were conducted under strict aseptic conditions. Equipment such as microscopes, incubators, centrifuges, refrigerators, water baths, autoclaves, anaerobic jars, and hot air ovens were checked regularly to ensure their proper functioning and the reliability of the results. The results were recorded in a neat and clear manner.

### 2.7. Statistical Analysis

The data was entered into a standard format computer database, checked for errors, and verified. Data maintained in the computer sheets were organized and analyzed using SPSS software (Version 21.6) and GraphPad Prism (Version 9.5.1). Data are presented in appropriate tables, figures, charts, and graphs by calculating percentages, rates, etc. Appropriate statistics were applied wherever applicable.

## 3. Results

Samples were collected from different biological specimens (pus, wound, swab, blood, urine, sputum, tissue, nasal swab, and ET) at Kathmandu Model Hospital, Kathmandu, Nepal. Of the 4,285 samples collected from the census, 848 (21%) samples had growth, and 3437 (79%) had no growth. Of the total samples, the male had a higher abundance of bacterial growth (Fig. 1)

**Fig 1:**
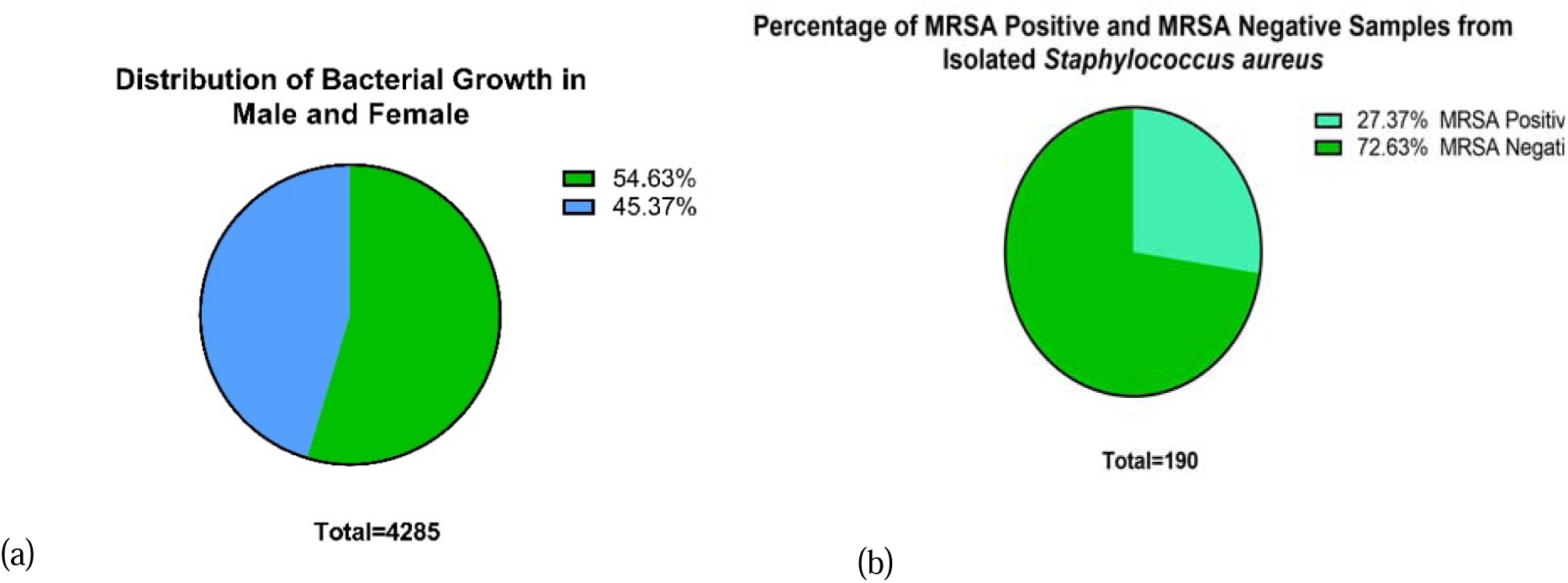
Distribution pattern of bacteria. Among the total samples of 4,285, 2341(54.63%) were male, and 1944 (45.36%) were female (a). Similarly, among 190 samples, 52 (27.37%) were Methicillin Resistant *Staphylococcus aureus* (MRSA), and the other 138 (72.63%) were MRSA negative (b).

Similarly, out of 848 growth of bacteria, 190 (22%) were *Staphylococcus aureus,* and the other 658(78%) were other bacteria. The isolated *Staphylococcus aureus* was further tested for MRSA for their antibiotic resistance. One-hundred ninety *Staphylococcus aureus* isolates were isolated from the clinical specimen taken during the study, among which 52(27.37%) isolates were MRSA.

Of the MRSA isolates, 29 (56%) were male, and 23(44%) were female from the total 52 MRSA samples. Males had a higher prevalence of MRSA than males (Fig 2). For further analysis, MRSA distribution was analyzed between the age of patients and the biological specimens. Among the tested antibiotics, vancomycin, teicoplanin, linezolid, and doxycycline were 100% resistant to MRSA isolates.

**Fig 2:**
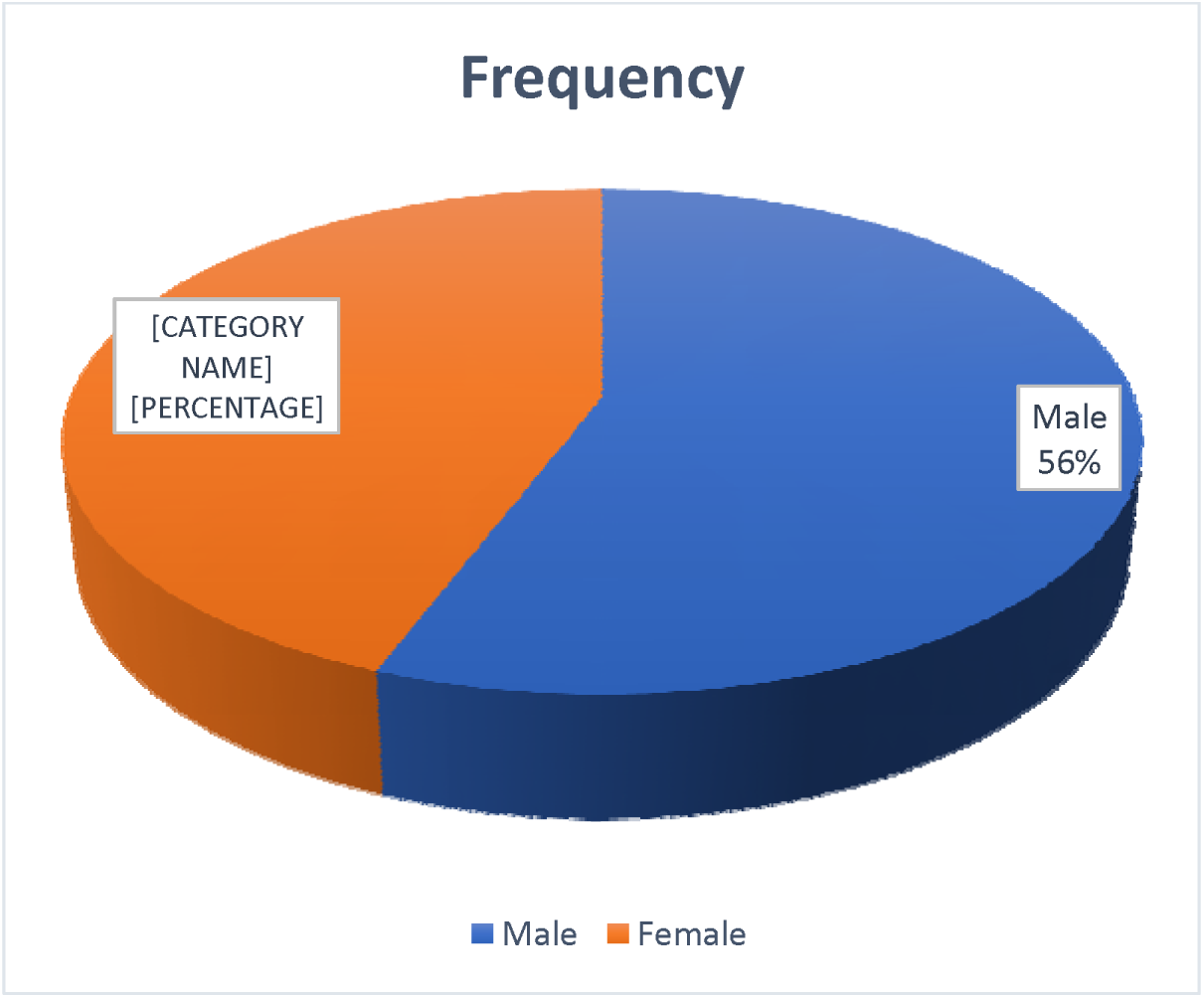
Number of positive MRS samples between males and females. Among the 52 MRSA-positive, 56% were male, and 44% were female.

The susceptibility pattern of the age group of participants had varying degrees of susceptibility to the antibiotics used, as shown in Fig 3. It was observed that as age progresses, the degree of resistance increases. Similarly, MRSA isolated from clinical specimens was found to be sensitive to different antibiotics used in this study. Compared to resistance, the specimen isolates were sensitive, as shown in Fig. 5.

**Fig 3:**
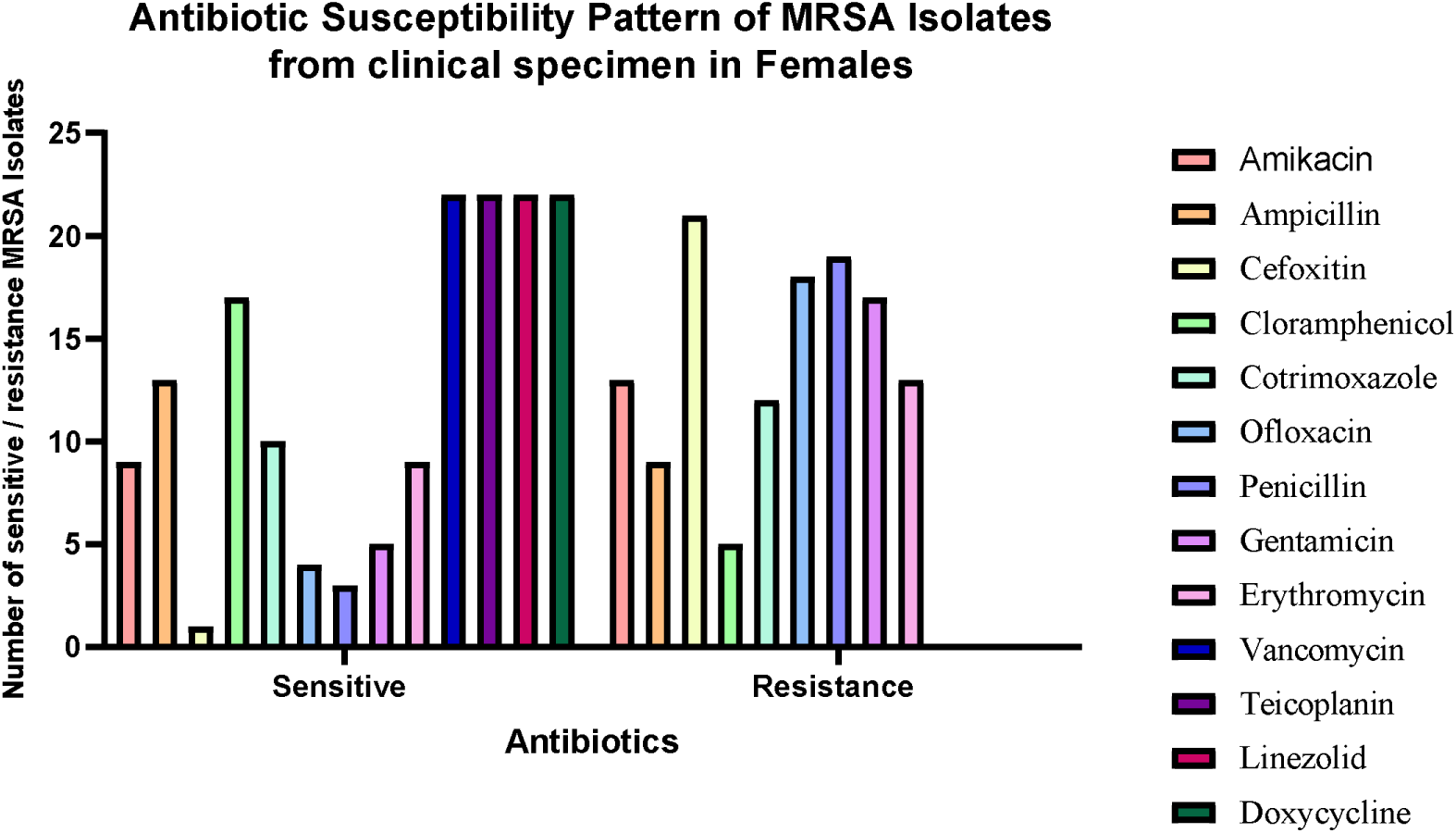
Antibiotic susceptibility pattern of MRSA Isolates from female clinical specimens. The higher female population was resistant to Cefoxitin and sensitive to Vancomycin, Teicoplanin, Linezolid, and Doxycycline.

**Fig 4:**
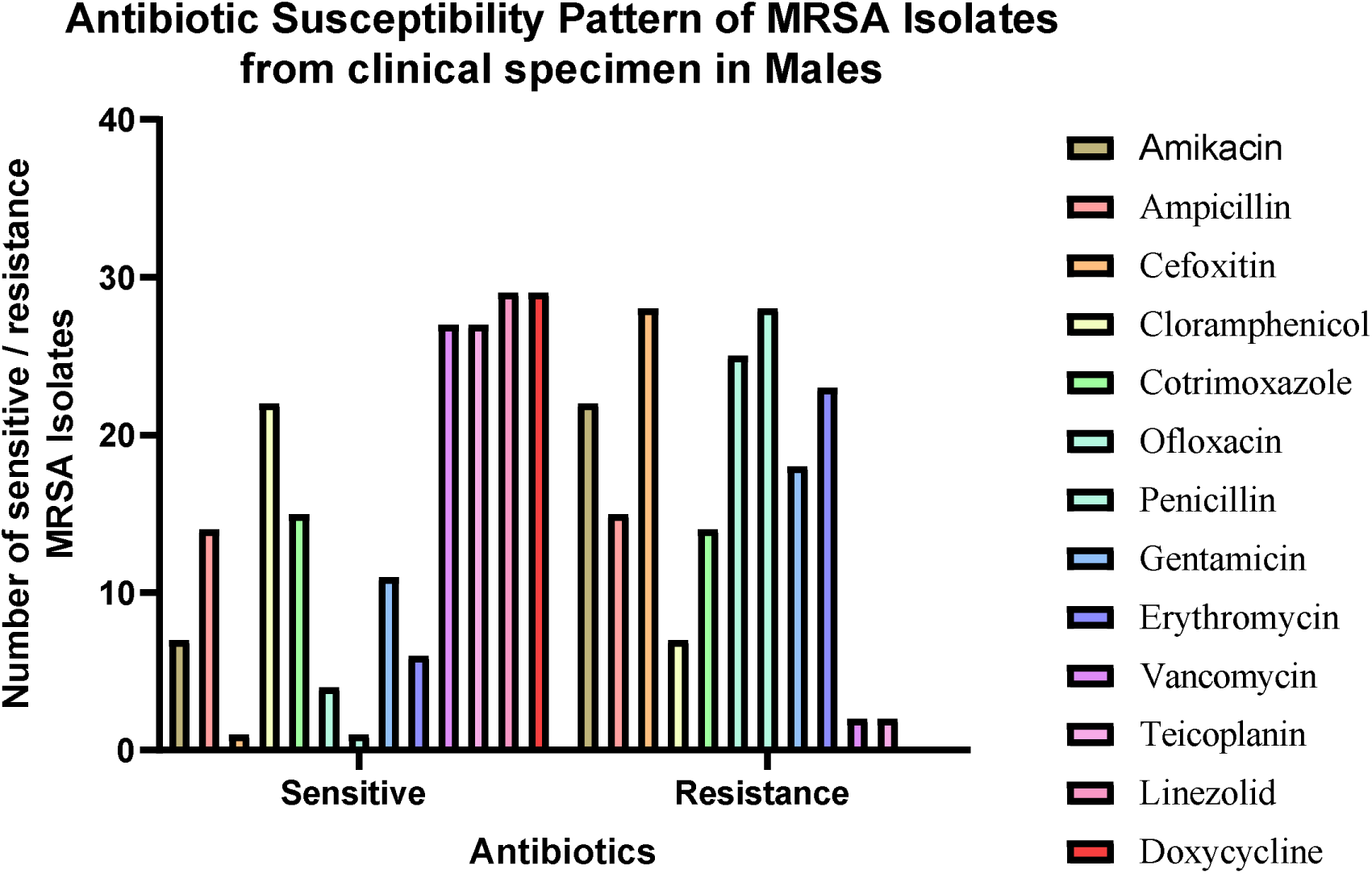
Antibiotic susceptibility pattern of MRSA Isolates from female clinical specimens. The higher male population was resistant to Cefoxitin and Penicillin and sensitive to Linezolid and Doxycycline.

**Fig 5:**
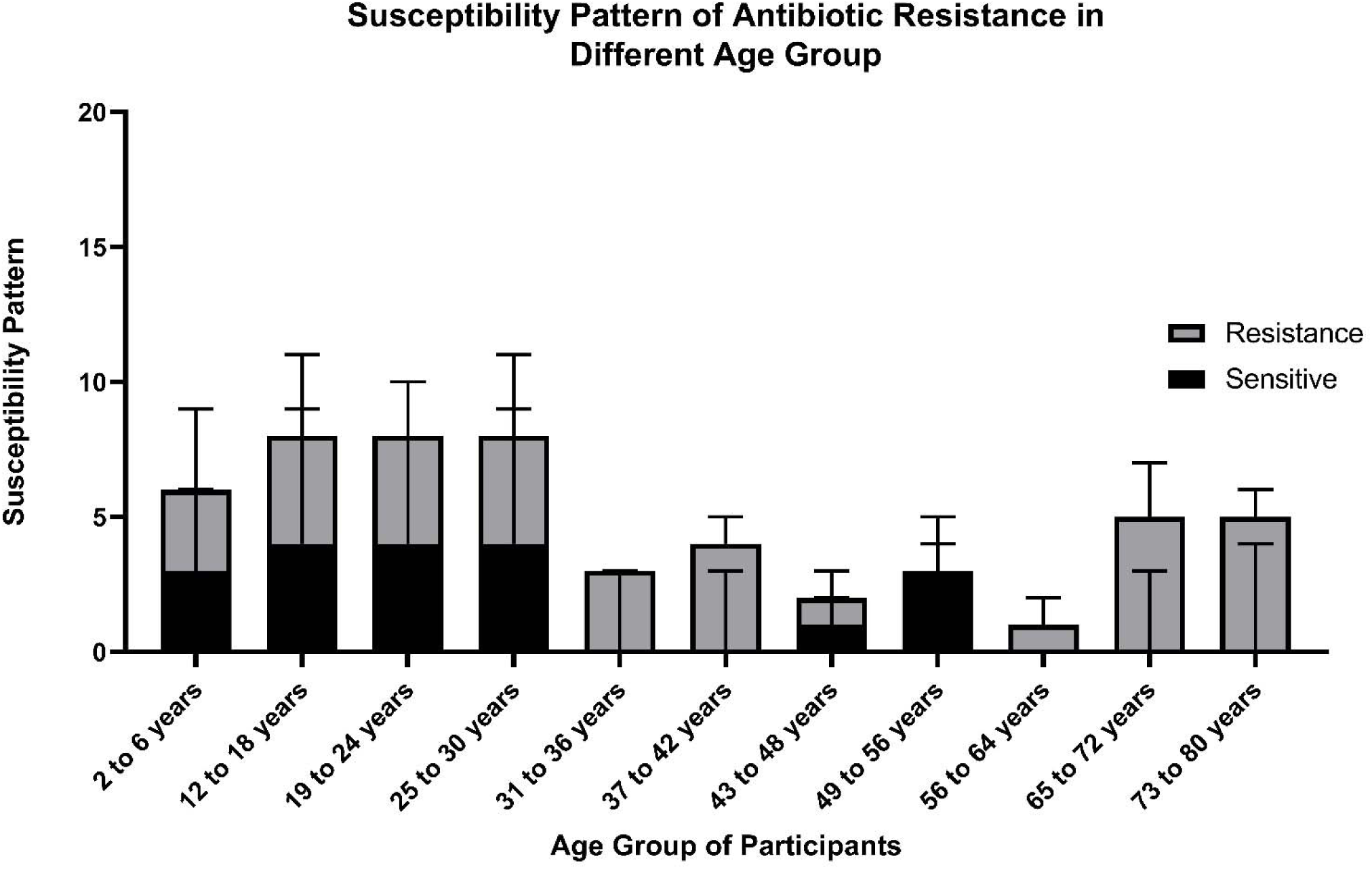
The study was carried out between age groups ranging from 2 years to 80 years patients. Other antibiotics disks were used to characterize the multi-drug resistance strains. Among the group of antibiotics used, the age group of 31-36 years, 37-42 years, and greater than 56 years were found to have a higher abundance of multi-drug resistance bacteria. The age group 49-56 years was found to be sensitive to the drugs used.

**Fig 6:**
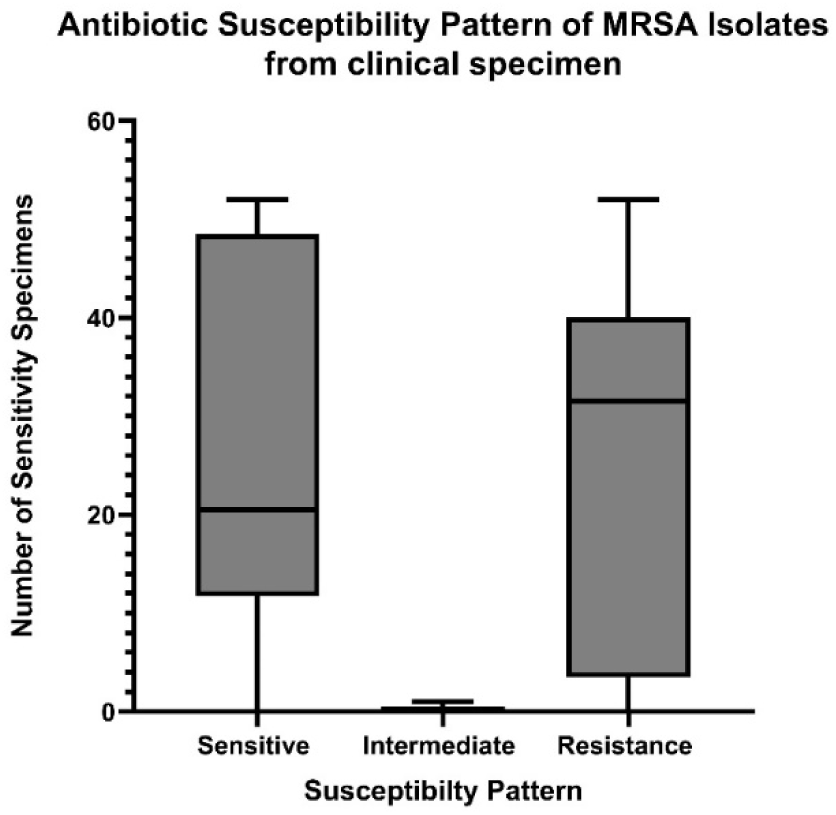
Higher number of clinical specimens were found to be sensitive to the antibiotics used in this study. A smaller number of samples showed intermediate effects, which were thus not further analyzed in this study. More emphasis was given to sensitivity and resistance patterns with higher expression levels. Large numbers of participants are from age 2-42 as opposed to the remaining numbers of age group 43-80. This might have led to a significantly higher proportion of specimens sensitive to antibiotics.

**Fig 7:**
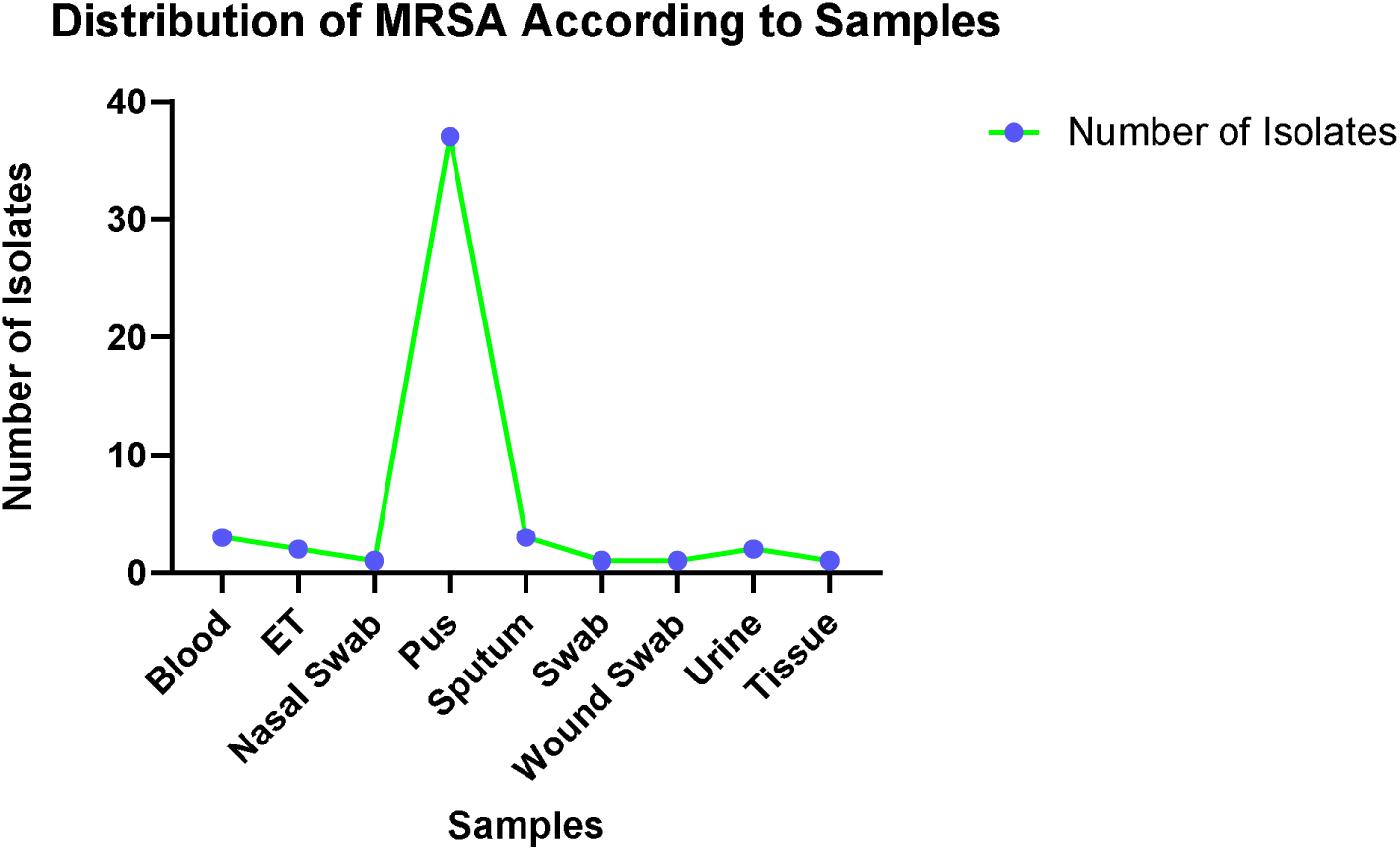
MRSA samples were abundantly present in the pus samples. MRSA was also significantly present in urine samples, which might be one of the leading causes of urinary tract infection in the patient. The total number of MRSA isolates in each specimen were: Pus −37, Blood-3, Ear and Throat (ET)-2, Nasal swab-1, sputum-3, wound swab-1, urine −2, and Tissue −1.

Among 52 clinical samples, more MRSA isolates were found in pus samples. There is a higher prevalence of MRSA in pus samples. Blood, urine, and ET were found to have an equal number of MRSA isolates.

Similarly, all the isolated MRSA isolates (52) showed 100% resistance against Penicillin and Cefoxitin, followed by Chloramphenicol (78.84%), Erythromycin (71.15%) and Amikacin (69.25%). Ampicillin and Co-trimoxazole showed the same level of resistance at 53.8%. MRSA was found to have the highest level of sensitivity toward Vancomycin. Almost all antibiotics except Erythromycin didn’t show any intermediate resistance.

**Table 1:** Antibiotic susceptibility pattern of MRSA isolates.

| Antibiotics | Sensitivity |  |  |
| --- | --- | --- | --- |
|  | Sensitive | Intermediate | Resistance |
| Amikacin | 16(30.7%) | 0 | 36(69.25%) |
| Ampicillin | 24(46.15%) | 0 | 28(53.8%) |
| Cefoxitin | 0 | 0 | 52(100%) |
| Chloramphenicol | 11(21.15%) | 0 | 41(78.84%) |
| Cotrimoxazole | 24(46.15%) | 0 | 28(53.8%) |
| Ofloxacin | 44(84.61%) | 0 | 8(15.39%) |
| Penicillin | 0 | 0 | 52(100%) |
| Gentamycin | 17(32.69%) | 0 | 35(67.3%) |
| Erythromycin | 14(26.9%) | 1(1.92%) | 37(71.15%) |
| Vancomycin | 50(96.15%) | 0 | 2(3.84%) |
| Teicoplanin | 52(100%) | 0 | 0 |
| Linezolid | 52(100%) | 0 | 0 |

PCR was done further to analyze the *<u>SSmec</u>* gene in isolated Hospital Acquired-MRSA strains. Results from PCR analysis revealed that out of 52 strains, 50 had the *mecA* gene (Fig. 9). Our study confirmed the presence of the *mecA* gene in 96% of strains, as shown in Fig. 8.

**Fig 8:**
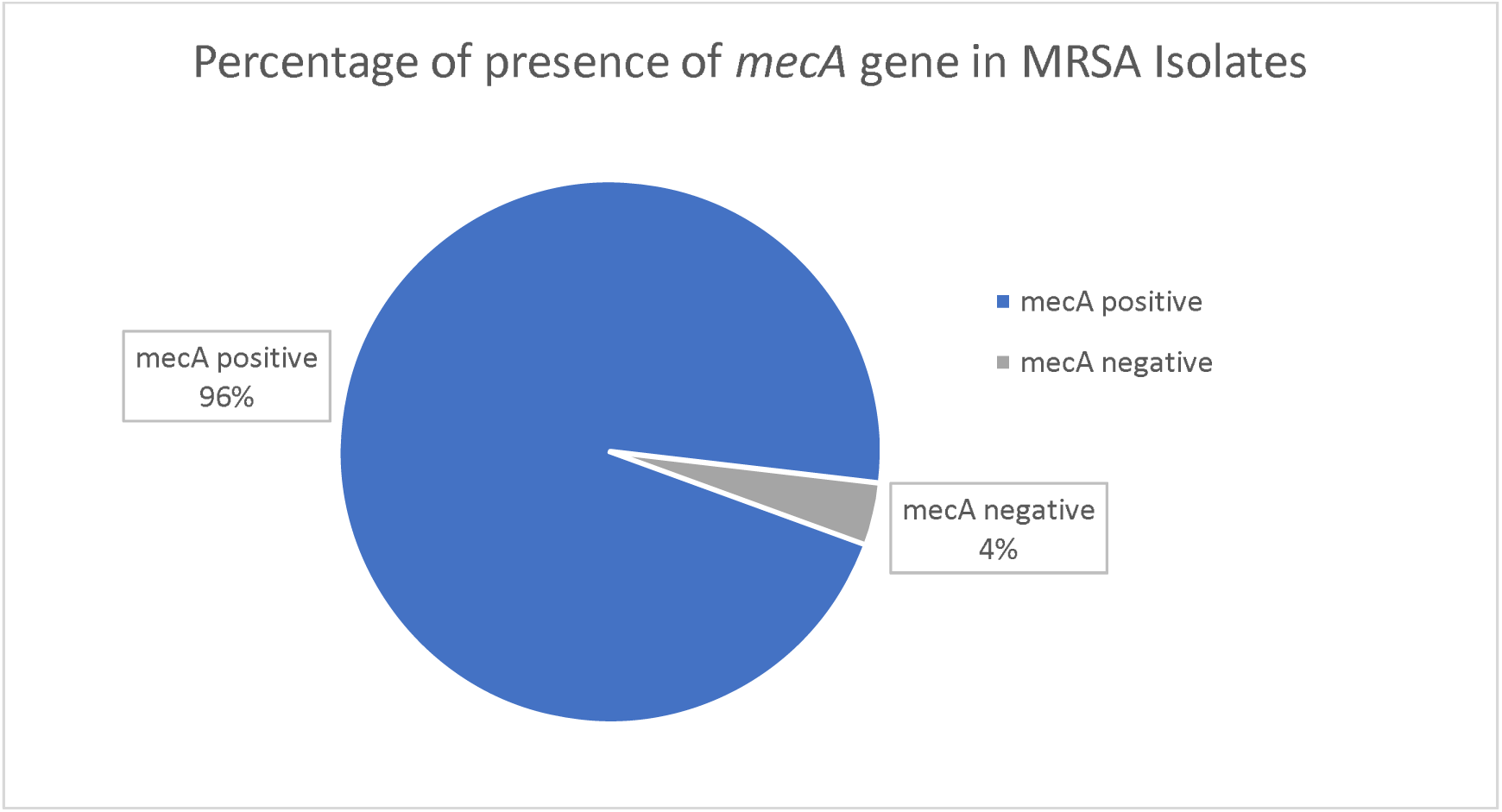
Out of 52 samples, 50 had positive *mecA* gene. Among the total MRSA isolates, *mecA* had a higher abundance than non-*mecA* positive samples.

**Fig 9:**
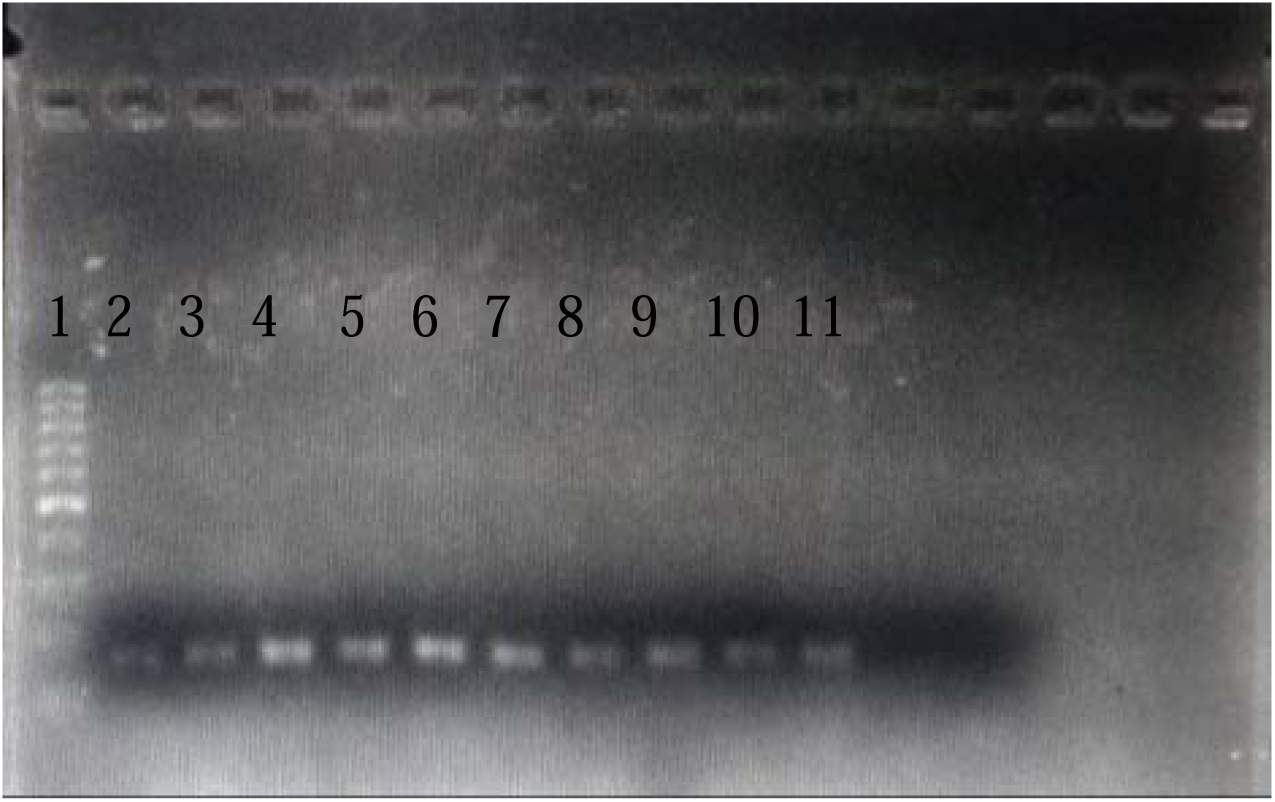
PCR product of *mecA* gene (163 bp): lane 1 100 bp ladder, lanes 2 to 11 positive samples for *mecA* gene. Only the positive samples with 163 bp are shown in the figure.

## 4. Discussions

Methicillin-resistant *Staphylococcus aureus* (MRSA) is a significant pathogen that has emerged in the last four decades, producing both nosocomial and community-acquired infections. Rapid and reliable diagnosis of methicillin resistance in *S. aureus* is critical for administering appropriate antimicrobial treatment and managing MRSA nosocomial dissemination [14].

The main aim of this study is to highlight the trend of the emergence of MRSA and detect the frequency of *mecA* in the isolated MRSA strains. Similarly, from this study, we could elucidate the resistance of MRSA to the empirical drugs used in hospitals and medical centers in Nepal.

One of the most significant discoveries in this investigation is the prevalence of MRSA strains (27.37%) among isolated *Staphylococcus aureus* from different biological specimens. MRSA strains were identified via the conventional method, the Kerby-Baeur disk method [15]. The incidence of MRSA in our study was higher in males than in females ( 56% vs 44%). This finding accords with the metanalysis conducted by Ghia CJ et al. 2020 where the male population across the studies accounted for 60.4% of cases as opposed to 39.6% in females [16]. Furthermore, 7-year long-term research by Kupfer et al. revealed that male gender was a significant risk factor for acquiring MRSA [17]. A higher abundance of MRSA in males has been attributed to behavioral practices that increase the rate of MRSA infections.

In addition to gender, we sought to analyze the distribution of MRSA between age groups. Our study found that the age group from 56 to 80 had all resistant MRSA strains, following the trend in other literature [18,19]. Since age has been regarded as an independent factor for MRSA incidence by the Center for Disease Control and Prevention (CDC) or the Robert Koch Institute, some things might be more consistent with the results [20]. However, older people have more risk factors that must be accounted for.

Compared to the previous studies by different authors, such as Pai V et al. 2020, we found a higher population of MRSA strains in the Pus sample (71%) [21]. In other investigations, pus was shown to have a more significant proportion of Methicillin-resistant *Staphylococcus aureus* than blood, urine, wound swab, and sputum [22,23].

Similarly, we analyzed the antibiogram of isolated MRSA from different clinical samples. All MRSA samples were found to be resistant to penicillin and cefoxitin. The resistance of MRSA to β-lactam antibiotics such as penicillin arises due to the mutation of penicillin-binding protein [24]. Chloramphenicol is effective against a large population of MRSA isolates worldwide [25]. But, in our study, we found 78.84% of strains of MRSA resistant to it. High resistance to chloramphenicol may be related to local antibiotic prescription habits. Resistance to erythromycin and gentamycin has rapidly developed. Due to the emerging resistance of MRSA to these classes, it has not been advised for the treatment of MRSA [26]. In this comparative analysis, vancomycin, teicoplanin, and linezolid were the most effective antibiotics. The synergistic actions of linezolid/vancomycin and linezolid/teicoplanin are effective against MRSA isolates [27]. Our results revealed and accorded with the superiority of linezolid and teicoplanin in treating MRSA, as evident in many studies worldwide [28, 29, 30]. Thus, this antibiogram of MRSA isolates shows that Teicoplanin and Linezolid, which belong to the glycopeptide and oxazolidinone classes, respectively, are the most effective antibiotics.

Almost 96% of the clinical samples in our study showed the *mecA* gene. The mecA gene’s high prevalence in MRSA indicates the potential resistance to the beta-lactam group. *mecA* gene is a gold standard for identifying MRSA [31]. Our results of the high *mecA* gene correlate with the study conducted by Haydeh E et al., 2019 and McTavish, SM et al., 2019 [32, 33]. In our study, only 4% didn’t have *mecA* gene. Some studies have reported a low occurrence of the *mecA* gene [34, 35]. The increased prevalence of the mecA gene in our study might be attributed to the fact that the samples were obtained from a regular diagnostic lab where there is a mixture of patients from the intensive care unit, extended stay of patients in the hospital, frequent use of invasive medical procedures, and haphazard use of multiple antibiotics [36].

Various intrinsic factors play a role in the development of resistance, suppressing the expression of the *mecA* gene. In a recent investigation, five major SCC *mec* types, *mecA,* and the PBP2 gene product were missing; the isolates remained phenotypically resistant, indicating the possibility of β-lactamase hyperproduction [37]. Similarly, another study revealed that specific changes in amino acids on protein binding cascades (PBPs 1,2 and 3), which play a crucial role in developing MRSA resistance, might alter the expression of *mecA* in resistant strains [38].

This study has a few limitations, including a shorter study duration, a smaller sample size, and a single study site. Future research might expand on the findings by doing a longitudinal study at several tertiary cares to enhance the results. Due to limited resources, minimum inhibitory concentration (MIC) was not possible, which might have shaded some further insights into this study. Nonetheless, this study emphasizes the phenotypic and molecular methods for detecting MRSA, providing a valuable reference for future studies on MRSA in Nepal and developing countries. Comprehensive information on the antibiogram of MRSA strains can be beneficial for tertiary centers where nosocomial infections are significantly high.

## 6. Ethical Approval

The ethical approval for this study was approved by the Institutional Review Committee (IRC) of the Public Health Concern Trust, Nepal (phect-NEPAL) to conduct the research project at phect-NEPAL/Kathmandu Model Hospital under IRC application number: 006-2020.

## 7. Consent

Written consent was obtained from all participants.

## 9. Authors’ Contributions

SN conceived the study and participated in the design. SN and AA wrote the manuscript and analyzed the data. SN performed laboratory experiments. AA performed the statistical analysis. SN and SS reviewed edited the final manuscript. All the authors read and approved the final manuscript.

## Data Availability

All data produced in the present study are available upon reasonable request to the authors

## Acknowledgments

The authors thank all the study participants, staff of the Kathmandu Model Hospital, and the Central Department of Biotechnology at Kirtipur, Nepal. The authors are grateful to the phect-NEPAL for the approval. We would like to express our deepest gratitude to the late Professor Dr. Manju Hada, Trichandra College, Kathmandu, Nepal whose invaluable guidance, expertise, and unwavering support were instrumental in the preparation and submission of this manuscript.

